# Integration of climate change and health components in medical education worldwide- a scoping review

**DOI:** 10.1101/2025.02.06.25321781

**Authors:** Basima Maisoon, Prakash Narayanan, Ranjitha S Shetty

**Affiliations:** Department of Global Health Policy and Governance, Prasanna School of Public Health, Manipal Academy of Higher Education, Manipal, Karnataka, India; Department of Community Medicine, Kasturba Medical College, Manipal, Manipal Academy of Higher Education, Manipal, Karnataka, India

## Abstract

There is growing recognition of the health impacts of climate change, prompting numerous calls and proposals in recent years for its inclusion in medical curricula. A scoping review was conducted according to JBI methodology to map and assess the integration of climate change and health content in undergraduate medical education globally and to identify existing curricula, educational strategies, and thematic coverage. Three scientific databases were searched for studies and articles that had instances of climate change integration in undergraduate medical curricula. The screening was done in stages following the PRISMA-ScR process, and there were 24 final inclusions. Data extracted include course type, duration, content, year of implementation, motive, challenges, enablers and recommendations. The review synthesized evidence from schools in North America, Europe, and Australia. There was a lack of studies from Low- and Middle-Income Countries (LMICs). The earliest instance of integration of climate change and health education was found to be in 2018, most schools started between 2020 and 2021. Common methods included stand-alone modules, electives, workshops, and modifying existing core curricula to include climate change and health components. Student-led initiatives significantly drove efforts in some schools. Courses emphasized climate change fundamentals, health impacts, vulnerabilities, communication and advocacy, interdisciplinary perspectives, and some practical applications. Challenges included integrating new content into existing curricula, requiring strategic planning and support. Recommendations include core curricular integration, experiential and practical learning, and encouraging research and advocacy.

## Introduction

Climate change (CC) refers to long-term changes in temperature, precipitation, wind patterns, and other aspects of the Earth’s climate system. The current climate change situation is anthropogenic in nature-primarily driven by human activities like the burning of fossil fuels, deforestation, and industrial processes.

CC can affect health adversely in multiple ways like causing heat related illnesses, weather-related injuries and fatalities, worsening chronic conditions, spread of vector borne diseases, infections from food and water, and mental health issues (1). With the growing recognition of health impacts of CC in recent years, there is an increasing awareness of the need to educate medical students on this subject. There have been calls worldwide for integrating climate change and health (CCH) and related disciplines like planetary health and environmental health, and sustainable healthcare into medical education (2, 3, 4, 5). Medical education bodies and councils have also recognized the need for inclusion of CCH in medical curricula. Some countries have taken the initiative to develop and modify graduate outcome statements and learning objectives of medical education that address CCH, as in the case of the Australian medical council (6). The General Medical Council outcomes for medical graduates in the U.K. recommend incorporating climate health and sustainability education into medical curricula, stating that newly qualified physicians must be able to integrate community health ideas, practices, and knowledge into their medical practice, as well as improve health and sustainable healthcare (7). Similarly, the American Medical Association approved a resolution in 2022 recognizing climate change as a public health emergency and stipulating advocacy for laws that facilitate the quick adoption of sustainable energy alternatives and climate resilience using a climate justice approach (8).

Given these developments, a scoping review would provide a comprehensive overview of how different countries and medical schools are integrating CCH into their curricula. This review would help identify gaps, best practices, and barriers to effective integration, thereby informing future educational policies and practices in medical education globally.

A preliminary search for existing scoping reviews on the topic was conducted on two databases and yielded no exclusive results. Hence, the scoping review was undertaken.

## Research question

The research question was: What is the extent and nature of literature on the integration of climate change and health components in medical education globally?

The objective of the research was to assess the extent to which climate change and health curriculum components are integrated into undergraduate medical education globally.

## Methods

### Study design

This study was a secondary research study, and the study design was a scoping review.

### Framework

This scoping review was based on the methodology given by the Joanna Briggs Institute (JBI) manual for Evidence Synthesis (9). Data extraction, analysis, summarising, and reporting were also done following JBI recommendations (10).

#### Population

Medical schools

#### Concept

Climate change and health inclusion in medical education

#### Context

Undergraduate medical education in all geographical regions and countries

### Search strategy

Use relevant search terms such as "climate change," “health,” and "medical education" and variations of these terms and Boolean operators (AND, OR) to refine search queries.

### Databases used

PubMed, Scopus, and Embase

### Keywords used

Climate change, climate

Health

Education, medical, medical school

### Search strings

#### Pubmed

(((climate) OR (climate change)) AND (health)) AND (((medical education) OR (medical school)) OR (medical curriculum))

#### Scopus

TITLE-ABS-KEY ( "climate change" ) OR TITLE-ABS-KEY ( "climate" ) AND TITLE-ABS-KEY ( "health" ) AND TITLE-ABS-KEY ( "medical education" ) OR TITLE-ABS-KEY ( "medical curriculum" ) OR TITLE-ABS-KEY ( "medical school" ) )

#### Embase

’climate’/exp OR ’climate’ OR ’climate change’/exp OR ’climate change’ AND ’health’/exp OR ’health’ AND ’medical education’/exp OR ’medical education’ OR ’medical school’/exp OR ’medical school’

The last date of the literature search was 31/01/2024.

### Selection of studies

The studies were selected based on the following criteria:

#### Inclusion Criteria

Peer-reviewed studies or articles on climate change education in undergraduate medical studies were considered. The review also considered scholarly perspectives, opinion articles, and commentary. Studies from any country or geographical region were included. All publications had to be in the English language. Articles with publication date between 2016 and 2023 were included.

#### Exclusion Criteria

Studies related to allied health sciences, public health, and postgraduate medical education were excluded. Articles that did not contain examples of implementations of climate change health (CCH) integration were not considered. Non-peer-reviewed articles, conference abstracts, grey literature, and review articles were also excluded.

### Screening of studies

Screening of studies was done in multiple stages. First, duplicates were removed.

Then, title screening and abstract screening were done, and relevant studies were included. This was followed by full-text screening. Data was extracted from the final included studies.

### Application used

Rayyan web application was used to remove duplicates and for title screening.

### Data charting

Data Charting was done in Google Sheets with shareable link.

#### Headers in the data extraction table

The headers in the data extraction table were:

- Year of study, institution, country.
- Year of course implementation
- Background/Rationale/Felt need to start the course
- Course characteristics-including target audience and type of course.
- Course contents
- Duration of the course
- Teaching methods
- Learning Objectives and Outcomes
- Enablers (internal & external)
- Challenges (internal & external
- Recommendations

### Collating, summarising, and reporting results

A descriptive quantitative analysis was done. Tables and figures were generated using MS Excel and Lucidchart. Baseline qualitative content analysis was done and the results were narratively summarized. JBI guidelines were followed (10), and the PRISMA-ScR (Preferred Reporting Items for Systematic Reviews and Meta-Analyses-Scoping Review) checklist was used to ensure completeness (S1 Table) (11).

## Results

The screening and inclusion of studies in the scoping review are illustrated using the PRISMA flow diagram for the scoping review process (12).

There was a total of 9873 records identified from three databases of which 3535 duplicates were removed. This was followed by the title and abstract screening. 46 reports were sought for retrieval for full-text screening, and 45 were screened. The final included studies were 24.

### Description of the included studies

There were 24 inclusions in the review which describe examples of CCH integration in medical schools from different countries. Out of these, two sets of studies pertain to CCH integration in two corresponding medical schools at different points in time. Thus, this review includes information about CCH integration in 22 medical schools.

### Findings from the studies

#### Countries

This review found that ten medical schools in the United States have integrated CCH components into their medical education, four in Germany, three in Australia, and one each in the United Kingdom, Norway, Netherlands, Canada, and Austria (Fig 2).

**Fig 1.**
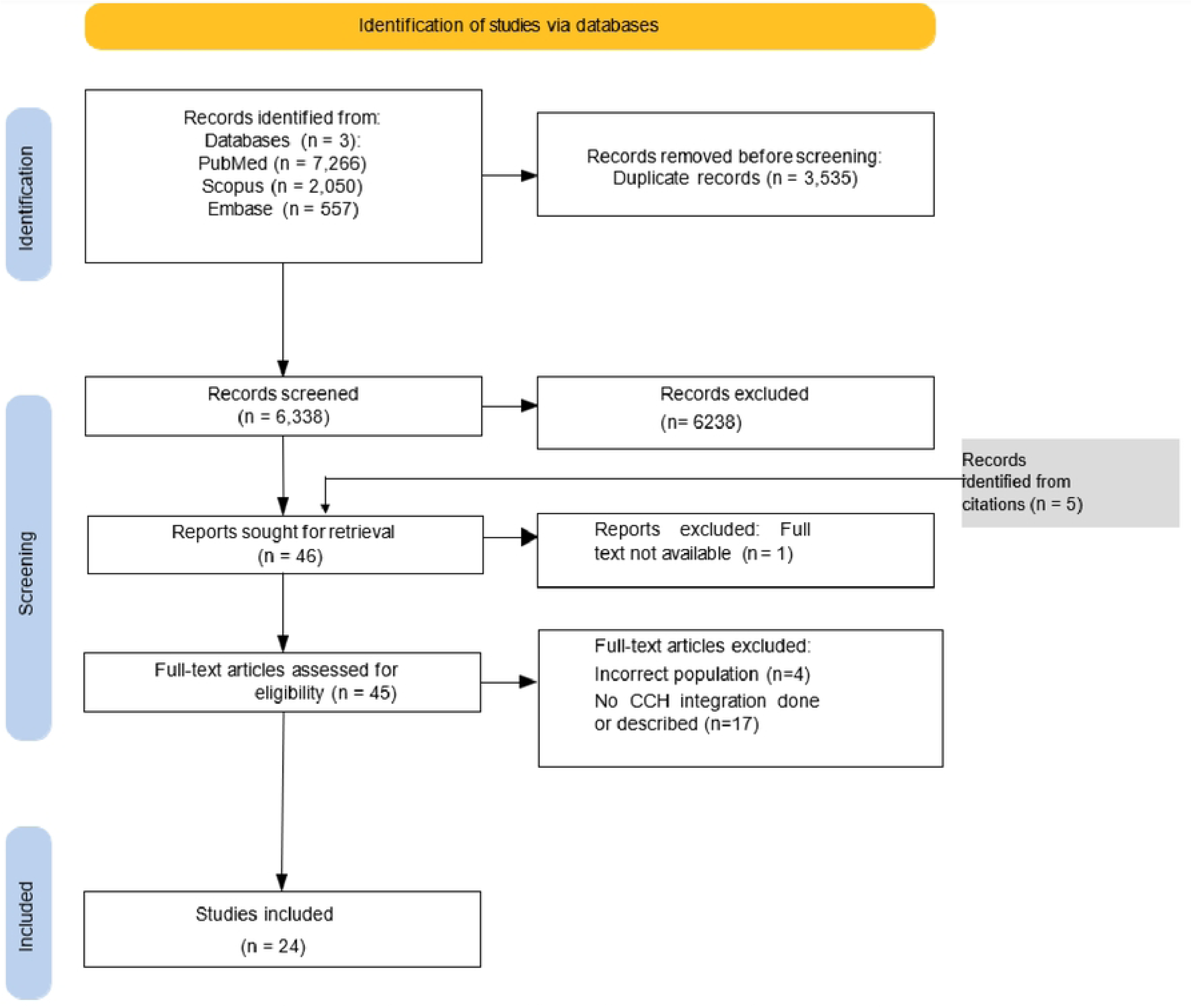
PRISMA flow diagram for the scoping review process.

**Fig 2.**
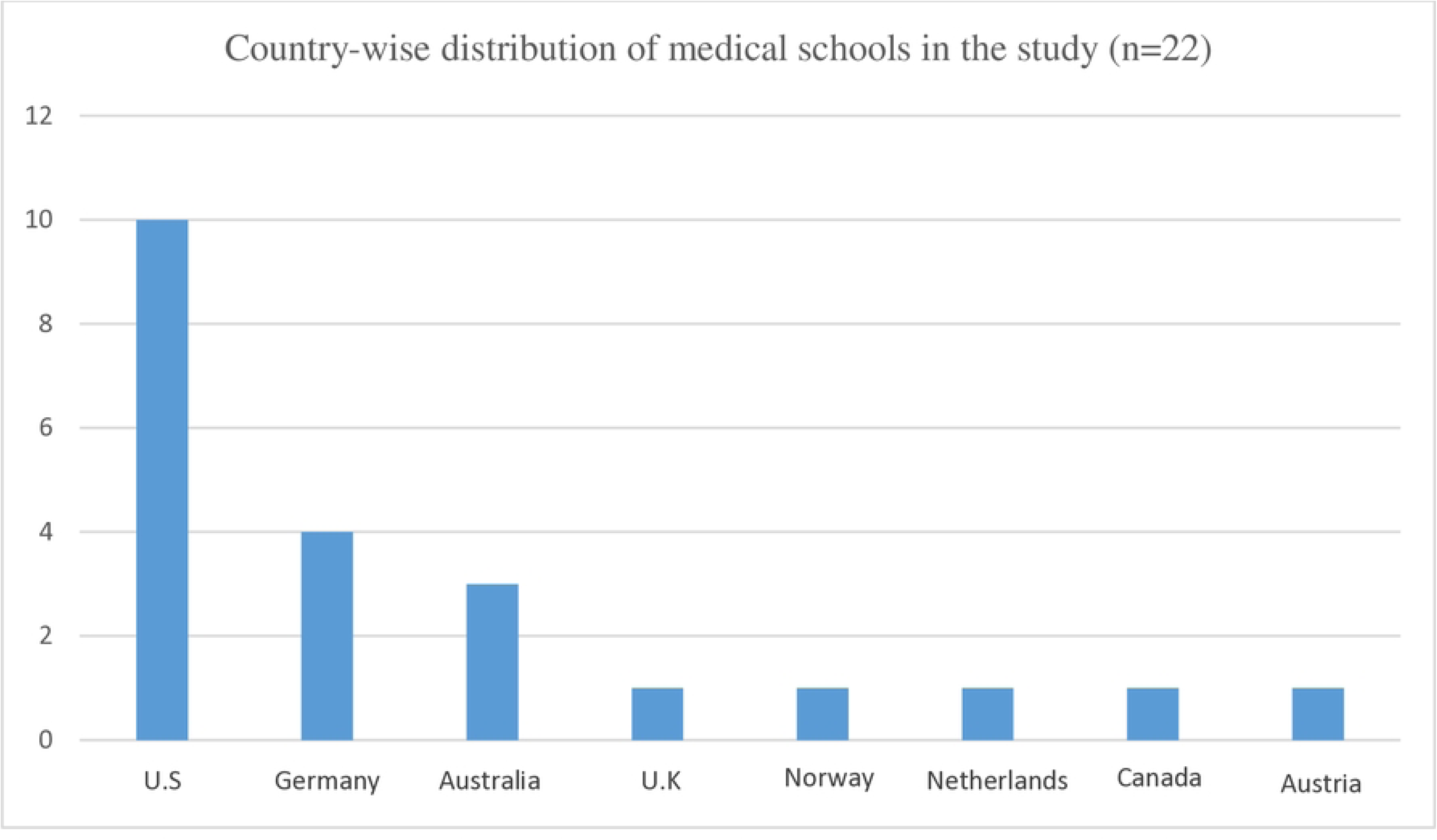
Country-wise distribution of medical schools that are teaching CCH.

#### Beginning year of integration

The earliest year of integration of CCH, as found in this review, is 2018. There are two instances of CCH integration in 2019, nine in 2020, seven in 2021, and three in 2022 (Fig 3).

**Fig 3.**
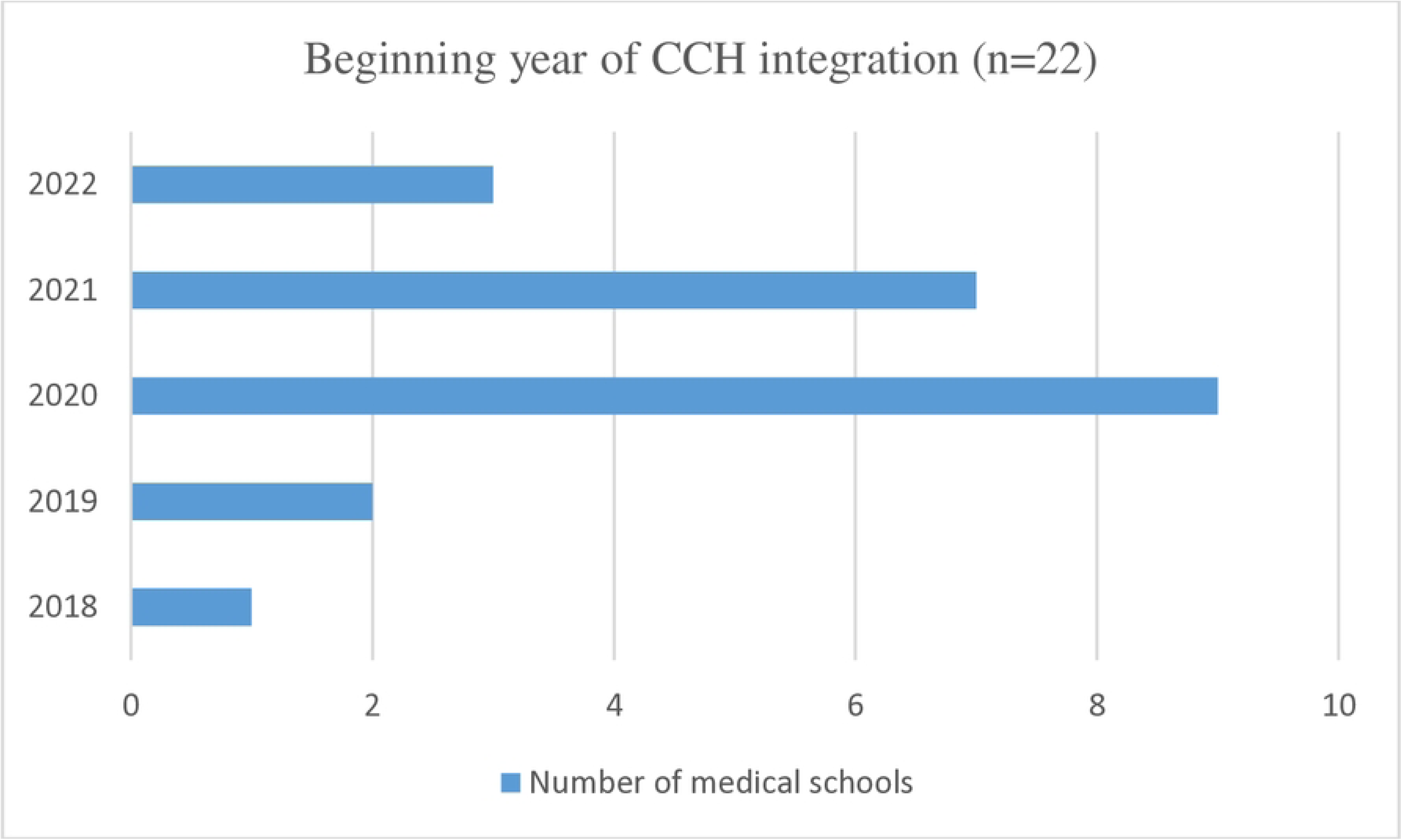
Distribution of the medical schools according to year of CCH integration (n=22) Target year of study and approaches to CCH integration.

Among the schools included in the review, it was found that CCH integration efforts were done in all years of study in four medical schools (14, 16, 26, 27) by integrating topics into the existing medical curriculum. The target group was students in clinical years of study in six medical schools (7, 17, 19, 20, 22, 23). Among these, four were electives on CC and two schools taught CCH as modules on planetary health.

CCH inclusion was targeted at preclinical years in eight schools (13, 15, 18, 21, 28, 30, 31, 34). Of these, two institutions implemented stand alone modules, while the remaining schools integrated CCH into the core curriculum. Notably, a unique climate change curriculum integration project was developed in a school (31); another focused on inquiry based learning with CC advocacy skill development and practical knowledge (30). Weekly links related to CC and planetary health were included in physiology lecture slides for six months in yet another school (21).

In some schools, workshops, conferences, seminars, or electives were conducted which were open to any year of study (8, 24, 25, 29).

Table 2 shows the different forms of integration.

**Table 1.** Country-wise distribution of sources include in the review (n=24)

| Country | Number of Sources |
| --- | --- |
| United States of America | 12 |
| Germany | 4 |
| Australia | 3 |
| United Kingdom | 1 |
| Norway | 1 |
| Netherlands | 1 |
| Canada | 1 |
| Austria | 1 |

**Table 2.** Approaches to CCH integration.

| CCH integration | Number of medical schools |
| --- | --- |
| Modified existing core curriculum | 9 |
| Standalone module | 11 |
| Workshop | 1 |
| Conference | 1 |

#### Motive

CCH integration in medical education was found to be student-led in eight medical schools (16, 18, 22, 24, 26, 27, 31, 33). Student advocacy led to creating a task force comprising medical students, faculty, and administrators, which led to curricular modification in one school(16). In another institution, student advocacy helped address the gaps in the curriculum regarding CCH and planetary health, and a course was co-created by students and faculty to address this (18). Similarly, a medical school was motivated by the already apparent effects of the climate issue and its relationship to the health sector, as well as by several students’ voluntary work, to start an elective (20).

In another school, an annual student-led conference on climate and health addressed the lack of formalized education on the health effects of climate change for medical students by offering a forum for interdisciplinary learning and discussion. The conference’s objectives included educating students about the urgency of climate change, its negative effects on human health, and how these health impacts disproportionately harm vulnerable populations (24).

In a medical institution in Canada, a country frequently affected by the adverse effects of climate change - medical students took part in advocacy campaigns and worked to improve education on the effects of climate change on health, which led to implementation of elective lecture series on the subject (26).

In an American medical school, the COVID-19 pandemic prompted medical students to reflect on a changing clinical situation. CC curriculum was not available to them throughout their preclinical years. Seeing the gap, they co-created a curriculum with the school curriculum committee, called "Climate Change & Environmental Health" which was taught to pre-clinical students (33).

In a Norwegian medical school, a course was designed in consultation with medical students with the goal of assisting students in gaining a nuanced understanding of CCH and the role of healthcare professionals (19). In another medical school in the US, student-led efforts were crucial in developing a unique climate change curriculum that was infused into core medical studies (31).

Medical institutions have taken the initiative themselves or followed the instructions of organizations and curricular bodies to begin the integration of CCH in many schools (7, 8, 13, 14, 15, 17, 19, 20, 21, 23, 25, 28, 29, 30). One school recognized that the medical curriculum only minimally addressed the subject of climate change in the past, with only a few stand-alone courses on the subject given. They saw a need for thorough and focused instruction and introduced an elective on the subject (13).

A medical school in the UK created an online module on climate change and sustainability in clinical practice, in order to meet learning objectives derived from the 2018 General Medical Council (GMC) graduate outcomes (7).

In another school, the 2022 resolution by the American Medical Association (AMA) declaring CC as a public health crisis and recognizing the role of physicians in climate advocacy prompted a medical school to conduct a workshop on CC (8).

An Australian medical school felt the need to find innovative ways of integrating important concepts in planetary health and climate change without adding to students’ workload and embedded CC concepts on Physiology slides while longitudinal planning for curriculum integration was underway (21).

Similarly, other medical schools felt the need for CCH integration in medical education, as it was identified as an educational priority (14) because CC is a social determinant of health (15), due to extreme weather events in their country (17), as part of teaching One health (20), and because healthcare providers must be sufficiently trained for clinical practice in the context of CC (23, 25, 28, 29).

#### Course contents

Review of course content on CCH revealed a range of topics. Fundamentals of climate change and its underlying science, health impacts of CC including physical and mental impacts were included in most schools.

##### Sustainable healthcare

Other key topics include impact of healthcare sector on climate (7, 13, 17), sustainable healthcare practices (7, 23), climate smart healthcare (25), sustainable development (19, 24), and a focus on Sustainable Development Goals (19). Education on infrastructure that promote sustainable behavior (30) and climate resilience in healthcare (29) were also imparted in some schools.

##### Biodiversity and Food security

Several institutions included the impact of CC on biodiversity (19, 28, 30) and food systems (19, 33) in their curricula. Additionally, the relationship between food security and CC was incorporated in some schools (21, 25, 26). Notably, one school implemented an urban garden project to teach students the links between nutrition, sustainable food practices, and CC (26).

##### Health equity

Topics related to health equity and justice were taught by many schools. This includes clinical ethics related to CC (13, 17), applying CCH knowledge to health equity (14, 16, 24, 25), and racial justice (16). CC education was also imparted by framing it as a social justice issue (31), environmental justice issue (33), and a social determinant of health (14, 29) highlighting disproportionate impacts.

##### Vulnerabilities

Vulnerabilities associated with CC, identifying climate vulnerable communities (24), education about population-specific risks in women’s health (16, 25), geriatrics, pediatrics (14), and in people with chronic disease conditions (16) were included in some CCH integration approaches.

##### Infectious diseases

Topics covered include the role of CC in infectious diseases (14, 20, 25, 31), its influence on disease patterns, and the importance of emergency preparedness to mitigate health risks (20, 25). The relationship between wildlife and public health was also addressed, highlighting the interconnectedness of ecosystems and human health (20).

##### Mitigation

Several schools integrated climate change mitigation education into their curricula. These included awareness of individual and professional roles (30), solutions at individual level like modifying own behaviour (15, 13), focus on the health co-benefits of climate mitigation strategies (7) etc. Solutions at political, technical levels (13, 22) and broader mitigation strategies were also taught (17, 26).

##### Communication

Communication regarding climate and health was covered in various programs. This included training in effective public health communication (15, 16, 19), patient consultations during climate crises (22), climate-sensitive health counseling (17) and learning about CC and practice of medicine (34).

##### Advocacy

Advocacy skills were prioritized in some schools, focusing on advocacy guidance (8, 16, 25) and training in designing effective advocacy campaigns (30).

##### Activities

Practical application of CC learning, like calculating ecological footprint (28), immersion learning experience on One Health (20), interactions and discussion with experts (22, 28), small group workshops (30), analysis of scientific publications (13), writing reflection papers (17), and assignments (23), were found.

#### Challenges and Enablers

Some challenges in CCH integration are that electives might have been done by those students who are already interested in the subject implying that other students miss the opportunity of learning about CCH (7, 17). Lack of assessment and evaluation of effectiveness were also challenges mentioned in some articles (17, 21, 32). Lack of time, less curriculum space, and scarcity of human resources were other challenges (8, 27, 30, 32). Lack of institutionalization and formal integration was mentioned as a challenge in a student-led effort (22).

Several strengths and enablers could also be identified from the studies. The time barrier was addressed by weaving CCH elements into existing content in some schools (14, 21, 31). Interdepartmental collaboration and involving multiple stakeholders were done to reduce barriers to curricular change and to increase available human resources (14, 20,24). An expert group was involved in a medical school (7), and consultation with students in another (19) to improve the efficacy of the course. Support of institutional leadership was mentioned as an enabler in some successful models (31, 34).

#### Recommendations

Recommendations from the studies include conducting mandatory courses on CCH as part of the medical curriculum and not just electives so that all students are familiar with the basics (8, 13, 22, 34). Further, many studies recommend incorporating CCH into existing core curricula and considering inclusion in the longitudinal curriculum (24, 25, 27, 30) or adding to formal teaching (26) to bring about sustained changes in student awareness regarding CCH. A study suggests integrating CCH into all levels of medical training (21). Continuous updation and integration of modules based on student feedback are recommended (7).

Diverse academic perspectives are to be included for a holistic learning experience (19). Other recommendations include teaching with experiential and group learning to improve students’ preparedness and self-efficacy (15), community-based and active learning opportunities (31, 34), ensuring research and advocacy opportunities (33, 34), including case simulations (16), and practical applications (7) in the curriculum.

Assessments to ensure a comprehensive evaluation of student knowledge on CCH were also recommended by some (18, 21,32).

## Discussion

The review includes insights about CC education in 22 medical schools from different countries. The inclusion of CC education spans across North America, Europe, and Australia, indicating a growing recognition of the importance of climate-related health issues. However, there were no studies from Low- and Middle-Income Countries (LMICs), suggesting a potential gap in these regions. The integration of CCH components in medical education seems to have begun relatively recently, with the earliest instance in this review in 2018. Most medical schools began integrating CCH components in their teaching from 2020-2021 onwards.

Different integration methods like stand alone modules on CCH, modifying core curriculum, workshops etc. were found in different schools.

The review reveals that in many medical schools, CCH integration efforts were driven by student-led initiatives. In some schools, courses on CCH have been co-created or developed after consultation with students. There is also an instance of an annual student-led conference that chose CC as the focus topic. This underscores the active role of students in advocating for the inclusion of climate change education, perhaps reflective of a grassroots movement within medical schools.

Many medical schools recognized CCH as an educational priority, acknowledging climate change as a significant social determinant of health that requires healthcare providers to be well-trained for clinical practice. Some were prompted by directives of medical councils and associations to teach CCH. The receptiveness to external guidelines, as well as internal advocacy, indicates a rising understanding of the significance of climate change education in medical education.

The research findings reveal varied approaches to CC education across various medical schools. Most schools covered climate change fundamentals, health impacts, and mitigation strategies. They emphasized interdisciplinary perspectives, integrating healthcare, environmental science, policy and advocacy to address the multifaceted challenges of climate change. The impact of CC on human-animal-environment interface, biodiversity loss, food insecurity, the intersectionality of CC and vulnerabilities were also included in CC education. There was focus on sustainable healthcare and topics such as the impact of the healthcare sector on climate, sustainable practices, and climate-smart healthcare were taught in many schools. This aligns with global efforts on sustainable development. Health co-benefits of climate mitigation and the promotion of climate-sensitive health communication were emphasised in some settings. Another notable feature is the integration of health equity and justice in CCH education. This imparts awareness of the disproportionality of impacts of CC and promotes critical thinking.

Practical components to CC education like growing urban garden, calculating carbon footprint, writing reflection papers and designing advocacy campaigns were identified in this review. Review and analysis of scientific publications on CC was also found. However, these were not widely implemented. Such approaches could enhance learning and create deeper understanding if more broadly applied.

The challenges identified include the practical difficulties of integrating new content into established medical curricula like lack of time and resources. This shows the need for strategic planning, resource allocation, and institutional support to ensure successful integration. The strengths and enablers point to effective strategies and best practices that can facilitate the integration of CCH. These include embedding content into existing courses, interdepartmental collaboration, leveraging external expertise when needed, student inputs, and institutional support.

The recommendations from the studies include core curricular integration, mandatory inclusion of CCH courses, incorporating diverse teaching methods and experiential learning that can enhance learning outcomes and practical skills. Assessment and feedback can ensure continuous improvement and relevance of the curriculum. Encouraging research and advocacy allows students to apply knowledge and contribute to the field. Implementing these recommendations could lead to a more robust and effective CCH education within medical schools, preparing future healthcare professionals to address issues more comprehensively.

## Conclusion

This scoping review has synthesized evidence on the integration of CCH education in medical schools globally. Medical schools are increasingly recognizing CCH as essential, with institutions adopting varied approaches to incorporate CC topics into their curricula.

There might be a great opportunity for institutions and curricular bodies to incorporate CCH components into medical teaching as well as to standardize CC education. By equipping students with necessary knowledge and skills, medical schools can play an important role in addressing CC.

### Limitations

This study could not include sources in languages other than English, potentially causing language bias. It is also possible that some relevant literature was missed or that the extraction of included sources may not have been thorough.

This study did not receive any funding.

## Data Availability

All relevant data are within the manuscript and its Supporting Information files

## Notes

### Competing Interest Statement

The authors have declared no competing interest.

### Funding Statement

The author(s) received no specific funding for this work.

